# Increased prevalence and clinical impact of hypocalcaemia in severe COVID-19 distinguishes it from other forms of infective pneumonia

**DOI:** 10.1101/2021.05.27.21257813

**Authors:** Meera R Mehta, Hakim Ghani, Felix Chua, Adrian Draper, Sam Calmonson, Meghna Prabhakar, Rijul Shah, Alessio Navarra, Tejal Vaghela, Andrew Barlow, Rama Vancheeswaran

## Abstract

**Background:** Hypocalcaemia has been reported in the context of acute COVID-19, where it has been associated with an increased risk of hospitalisation and disease severity. Calcium is an important intracellular messenger that controls diverse cellular processes. Two other clinically important coronaviruses, SARS-CoV-1 and Middle East respiratory syndrome (MERS)-CoV, can use calcium ions to enter and replicate within host cells. Calcium may therefore be important in the pathophysiology of COVID-19 infection. We sought to investigate whether calcium derangement was a specific feature of COVID-19 that distinguishes it from other infective pneumonias, and its association with disease severity.

**Methods:** We conducted a single centre retrospective study of albumin-corrected serum calcium on adult patients with COVID-19 who presented between March 1^st^ and May 16^th^ 2020. The primary outcome was maximal level of care based on the World Health Organization Clinical Progression Scale for COVID-19. Cases with community acquired pneumonia (CAP) and viral pneumonia (VP) were identified through a clinical database over three intervals (January to February 2018, January to February 2019 and September to December 2019).

**Results:** We analysed data from 506 patients with COVID-19, 95 patients with CAP and 152 patients with VP. Hypocalcaemia (serum calcium <2.2mmol/L) was a specific and common clinical finding in patients with COVID-19 that was not present in other respiratory infections. Calcium levels were significantly lower in those with severe disease. Ordinal regression of risk estimates for categorised care levels showed that baseline hypocalcaemia was incrementally associated with odds ratio of 2.33 for higher level of care, superior to other variables that have previously been shown to predict worse COVID-19 outcome. Serial calcium levels showed improvement by day 7-9 of admission, only in in survivors of COVID-19.

**Conclusion:** Hypocalcaemia may independently predict not only more severe but more progressive disease and warrants detailed prognostic investigation. The fact that decreased serum calcium is observed at the time of clinical presentation in COVID-19, but not other infective pneumonias, suggests that its early derangement is pathophysiological and may influence the deleterious evolution of this disease. If calcium is ultimately shown to be critical to the entry and replication of SARS-CoV-2 in host cells, unravelling how this mechanism could be therapeutically targeted deserves more intensive examination.

**Trial registration HRA:** 20/HRA/2344.

## Introduction

COVID-19, the disease responsible for the pandemic caused by the severe acute respiratory syndrome coronavirus-2 (SARS-CoV-2), has resulted in nearly three million deaths worldwide. The identification of well-defined COVID-19 prognostic markers has advanced the clinical understanding of this disease (1–3). Hypocalcaemia has been reported in the context of acute COVID-19 where it has been associated with an increased risk of hospitalisation and morbidity (4–9). However, its role in predicting mortality has been less clear, with reports of a conflicting effect (6,10–12)

Neither the pathogenic role played by hypocalcaemia in COVID-19 nor the possibility that its occurrence might simply reflect underlying illness severity has been established. However, the fact that decreased serum calcium is consistently observed at the time of clinical presentation suggests that its early derangement may potentially influence the deleterious evolution of this disease at an individual level.

Calcium is a versatile intracellular messenger involved in controlling diverse biological processes spanning gene expression, cell proliferation and cell death (13). Its role in promoting pathogen apoptosis during intracellular infection has been recognised in host defence (13). Conversely, some viruses have evolved ways to promote calcium availability to their own pathogenic, replicative and survival advantage (14–16). More specifically, alterations in calcium homeostasis effected by enhanced ion transportation through coronavirus envelope (E) proteins that act as viroporins has been linked to the activation of inflammatory cascades by the SARS-1 coronavirus (17). It has recently been proposed SARS-CoV-2 may also utilise this or similar mechanisms to augment its infectivity during host-virus interactions (18)

In the current retrospective study, we compared the prevalence of hypocalcaemia and its association with clinical outcome in different forms of infective pneumonia including COVID-19. We found an increased frequency of hypocalcaemia in COVID-19 that distinguished it from the other pneumonias and observed a correlation between low calcium and increased COVID-19 severity as assessed by indices of oxygenation, leucocytosis and extent of chest radiographic abnormality. Further investigations revealed an association between hypocalcaemia and increasing levels of respiratory support in the sickest patients with COVID-19. Moreover, in observing that low calcium levels subsequently improved in survivors of COVID-19, we postulated that hypocalcaemia may independently predict not only more severe but more progressive disease. In this regard, its role in COVID-19 prognostication warrants detailed prospective investigation.

## Methods

Adult patients aged 18 years or older with positive SARS-CoV-2 nucleic acid testing by real-time polymerase chain reaction (rRT-PCR) were retrospectively recruited at Watford General Hospital, West Hertfordshire NHS Trust between March 1^st^ and May 16^th^ 2020 (NHS HRA: 20/HRA/2344). A pre-specified study protocol including data collection methodology has been published elsewhere (19). Control cases diagnosed with community acquired pneumonia (CAP) and viral pneumonia (VP) were identified through a clinical database for three defined intervals over two preceding winters prior to the emergence of COVID-19 (January – February 2018, January – February 2019 and September – December 2019).

CAP was diagnosed on the basis of respiratory symptoms and signs indicative of a lower respiratory tract infection, with new plain radiographic abnormalities including lobar infiltrates or consolidation (20). VP was diagnosed by positive viral identification in swabbed nasal and oropharyngeal acute respiratory specimens and analysed by the BioFire® respiratory panel, which identifies amongst others *Influenzae A* and *B, Rhinoenterovirus, Human Metapneumovirus, Coronaviruses* (OC43, HKU1, NL63), *Picornovirus* and *Respiratory Syncytial Virus*.

An analysis was first undertaken to highlight differences in core clinical characteristics between patients with COVID-19, CAP and VP. Subsequently and only for the COVID-19 subgroup, the impact of an abnormally low calcium level on the maximum level of hospital care, as a surrogate of COVID-19 severity, was evaluated. The selection of maximal care level as the primary outcome was based on the World Health Organization (WHO) Clinical Progression Scale for COVID-19 (21). Patients were categorised at four levels of care: (i) mild ambulatory disease qualifying for monitoring in a virtual hospital (VH) setting, (ii) moderate hospitalised disease for individuals managed on a medical ward who intermittently required supplemental oxygen, (iii) severe hospitalised disease for patients who received continuous positive airway pressure (CPAP) therapy, and (iv) severe hospitalised disease who required invasive mechanical ventilation (IMV) on the intensive care unit (ICU).

Hypocalcaemia was defined as albumin-corrected serum calcium <2.2 mmol/L in serum, a level based on study-defined cut-points. An optimal cut-point of 2.195 mmol/L was associated with a sensitivity of 47%, specificity 43% and univariate odds ratio of 1.51 for in-hospital death (95% CI 1.14 – 2.00; p<0.01). We excluded patients with chronic kidney disease (22) and those receiving vitamin D or calcium supplementation. An index of oxygenation was calculated using the oxygen saturation-to-fraction of inspired oxygen (S:F) ratio, determined by dividing the SpO_2_ by the FiO_2_. A cut off of 4.38 represented saturations of 92% on fraction of inspired oxygen 21% (room air).

A subgroup analysis was undertaken to assess 25 Hydroxy Vitamin D level in patients with COVID-19 pneumonia; because vitamin D was not routinely measured as part of protocolized COVID-19 care, we included the result of a small subgroup of patients with mild or severe COVID-19 (presenting between December 2020 and January 2021) where the measurement was recorded prospectively within three days of a positive SARS-CoV-2 rRT-PCR test.

### Statistical analysis

Continuous variables were presented as median with interquartile range (IQR) and the Kruskal-Wallis equality-of-population or H test was used to compare differences among groups. Categorical variables were expressed as frequency (%) and analysed using Pearson’s χ^2^ test or Fisher’s exact test. To assess the effect of hypocalcaemia on the primary outcome, and taking account of potential co-variate effects with other independent variables, we used ordinal (ordered) logistic regression to compare the odds ratios of these variables on pre-defined outcome categories of maximal care level. A goodness-of-fit (Brant) test was employed to ensure that the proportional odds assumption was met. A non-significant test statistic (P>0.05) provided evidence that the full model did not show a significant difference to restricted models with selected explanatory variables. Following this, a partial proportional odds model was refined to test parallel odds against alternatives (Peterson and Harrell, 1990). A two-sided p value of <0.05 was considered statistically significant. All analyses were performed using GraphPad Prism V.9.0.2 and STATA, V.16 (Stata, Texas, USA).

## Results

### Differences in demographic and clinical characteristics between study subgroups

We analysed data from 506 patients (54.9% male) with COVID-19, 95 patients with CAP (53.6% male) and 152 patients with VP (44% male). The median age of the COVID-19 cohort was 65 (IQR 52-80) similar to that of VP patients (68, IQR 45-79) but lower than the median age of patients with CAP (79; IQR 67-86). **(Table 1)**

**Table 1.** Baseline Characteristics of patients with COVID-19 and non-COVID-19 pneumonia.

|  |  | **COVID-19** | **CAP** | **VP** |
| --- | --- | --- | --- | --- |
| **Total (N)** |  | **506** | **95** | **152** |
| **Median Age (IQR)** |  | 65 (52-80) | 79 (67-86) | 68 (45-79) |
| **Gender (n; % of N)** | Male | 278 (54.9%) | 51 (53.6%) | 67 (44.0%) |
|  | Female | 228 (45.0%) | 44 (46.3%) | 85 (55.9%) |
| **Ethnicity (n; % of N)** | White | 383 (75.6%) | 81 (85.2%)* | 134 (88.1%)* |
|  | Asian | 83 (16.4%) | 11 (11.5%)* | 12 (7.8%)* |
|  | Black | 30 (5.9%) | 1 (1.0%)* | 0 (0%)* |
|  | Other | 10 (1.9%) | 2 (2.1%) | 6 (3.9%) |
| **Smoking (n; % of N)** | Never | 420 (83%) | 75 (78.1%) | not known |
|  | Ever | 86 (16.9%) | 21 (21.8%) | not known |
| **S:F ratio <4.38** |  | 191/401 (47%) | 40/78 (51%) | not known |
| **RR >24 breaths/min** |  | 235/455 (51%) | 47/87 (54%) | not known |
| **CRP > 50mg/L** |  | 304/501 (60%) | 63/94 (67%) | 68/151(45%) |
| **WCC >11 X10<sup>9</sup>/L** |  | 96/505(19%) | 56/95 (58%)* | 92/151 (60%)* |
| **Lymph <0.7 X10<sup>9</sup>/L** |  | 251/505 (55%) | 56/95 (58%) | 48/151 (31%) |
| **CXR ≥4 abnormal zones** |  | 224/487 (45%) | 5/95 (5.2%)* | 4/147 (2.7%)* |
| **Median LOS (days)** |  | 7.5 | 5.43 | 4 |
| **Mortality (%)** |  | 30.60% | 28% | 4.60%* |

Baseline characteristics of patients with COVID-19, Community Acquired Pneumonia (CAP) and Viral Pneumonia (VP) compared. S:F ratio= saturations: fraction of inspired oxygen ratio, RR= respiratory rate, WCC= White cell count, Lymph= lymphocyte count, LOS= length of stay. \*p value < 0.05 versus COVID-19

Amongst patients with COVID-19, individuals of White Caucasian ethnicity constituted 75.6% (383/506) of the cohort while Asian, Black and other minor ethnicities (BAME) represented 16.5%, 4.5% and 1.7%, respectively. In comparison, White Caucasian ethnicity comprised a higher proportion of patients with CAP (85.2%) and VP (88.1%) (p<0.001 vs COVID-19).

To assess the level of severity of pneumonic illness across the three cohorts, we used a panel of common clinical variables conventionally taken to indicate disease severity at presentation. We found similar levels of hypoxia, tachypnoea and baseline CRP in patients with COVID-19 and CAP. Whereas CAP was associated with higher peripheral leucocytosis, a significantly higher proportion of patients with COVID-19 had multifocal infiltrates affecting 4 or more zones on plain chest radiography. Patients with VP had significantly lower CRP and limited radiographic changes.

The median length of hospital stay in patients with COVID-19 was significantly longer compared with CAP and VP: COVID-19 7.5 days (IQR 4.0-13.2), CAP 5.4 days (IQR 2.2-10.5) and VP 4.0 days (IQR 1.1-9.5) (p < 0.01). The in-patient mortality amongst COVID-19 patients was 30.6%, similar to that of patients with CAP (28%) but significantly higher than patients with viral pneumonia (4.6%, p<0.0001 COVID-19 vs. VP) (**Table 1**).

Hypocalcaemia was evident in 53% of patients with COVID-19 at initial assessment but only in 11.5% (11/95) and 10.5% (16/152) of patients with CAP and VP respectively (p<0.0001 vs COVID-19) (**Figure 1**). Hypocalcaemic patients with COVID-19 were more likely to be male (62.1% vs. 46.0%; p<0.001) and obese (34.0% vs. 23.8%, p<0.05). Median corrected calcium in COVID-19 was 2.19 mmol/L (IQR 2.11-2.27) compared to 2.31 mmol/L (IQR 2.24-2.38) in patients with CAP and mmol/L (IQR 2.24-2.44) in those with VP. The between-group difference in baseline serum adjusted calcium was significant (p<0.001).

**Figure 1.**
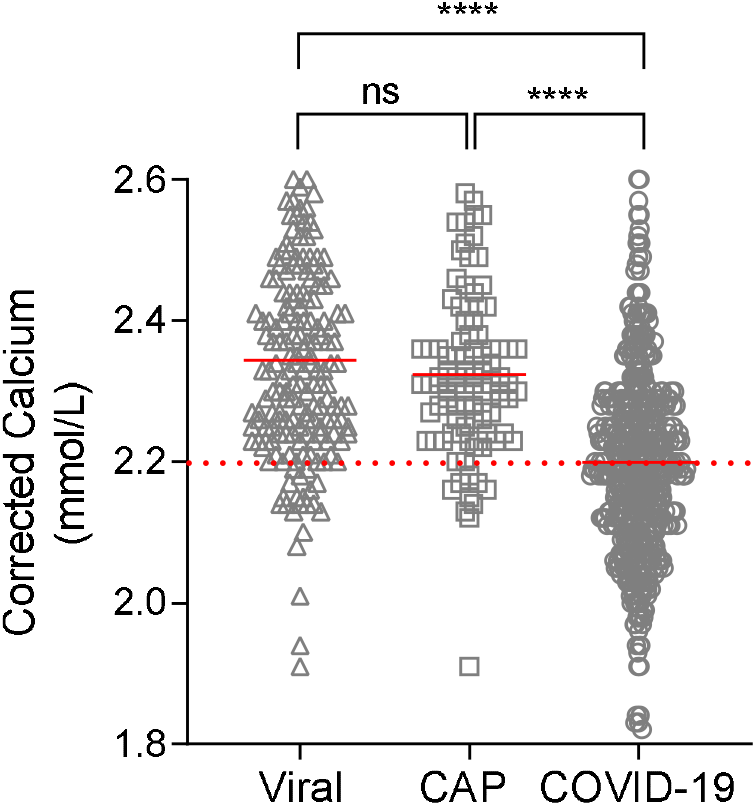
Serum Corrected Calcium at presentation in patients with Community Acquired Pneumonia (CAP), Viral Pneumonia (VP) and COVID-19. Dashed line represents cut-off point for hypocalcaemia, Solid red line represents median, ns= not significant, **** p <0.0001

### Association of hypocalcaemia with indices of infection severity in COVID-19

Hypocalcaemia in patients with COVID-19 was associated with greater physiological, biochemical and radiological derangement than normocalcaemia (**Figure 2**). Crucially, hypocalcaemic patients had a higher median respiratory rate compared to normocalcaemic patients (24/min, IQR 20-30 vs 22/min, IQR 18-28; p<0.005) and a lower baseline S:F ratio indicative of worse hypoxia at presentation (4.38 vs 4.52; p<0.01). The extent of baseline radiographic abnormality attributed to COVID-19 pneumonia was similarly greater in those with hypocalcaemia (p<0.001 vs normocalcaemia). Furthermore, hypocalcaemia was associated with a higher CRP (85, IQR 42-148 vs 60, IQR 32 -124; p<0.01). Although peripheral leucocytosis was not associated with hypocalcaemia, patients with low calcium had statistically worse lymphopenia than normocalcaemic individuals even though the difference was small (0.85 x10^9^/L vs 0.98 × 10^9^/L; p<0.05).

**Figure 2.**
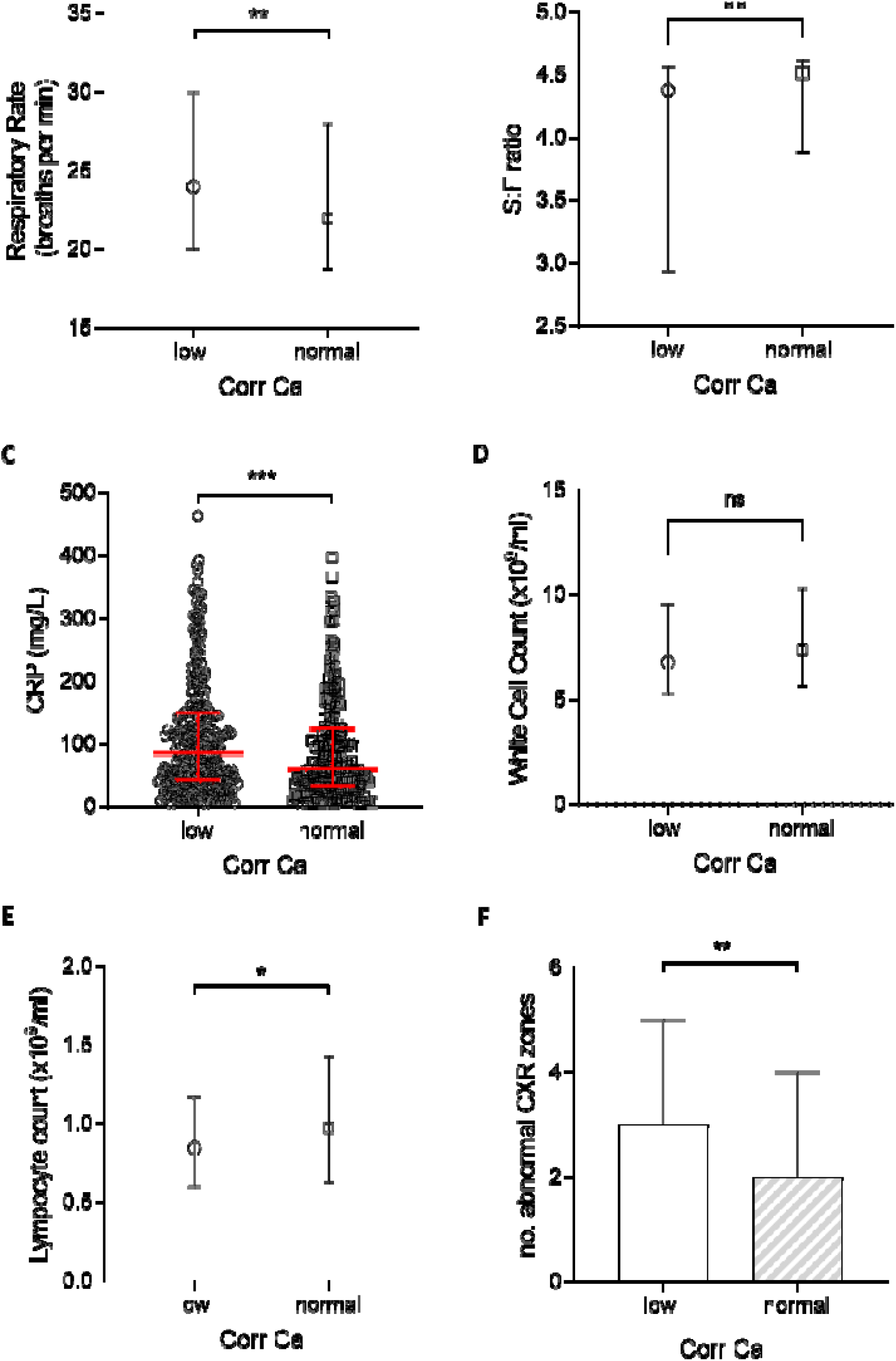
Physiological, biological and radiological severity markers in hypocalcaemic (corrected calcium < 2.2mmol/L) and normocalcaemic (≥ 2.2mmol/L) COVID-19 patients. (A) Respiratory Rate (B) oxygen saturation: fraction of inspired O2 (S:F) ratio (C) White Cell count (D) C-Reactive Protein (E) Lymphocyte Count and (F) number of abnormal chest X-ray (CXR) zones. Plots show median and IQR, ns= not significant, * p< 0.05, ** p<0.01 *** p< 0.001

### Hypocalcaemia as a predictor of higher levels of respiratory care in patients with COVID-19

Median serum calcium level was lower in those with moderate disease compared to those with mild COVID-19 (p<0.001), and lower still in those with severe disease (p<0.0001) (**Figure 3A**).

**Figure 3.**
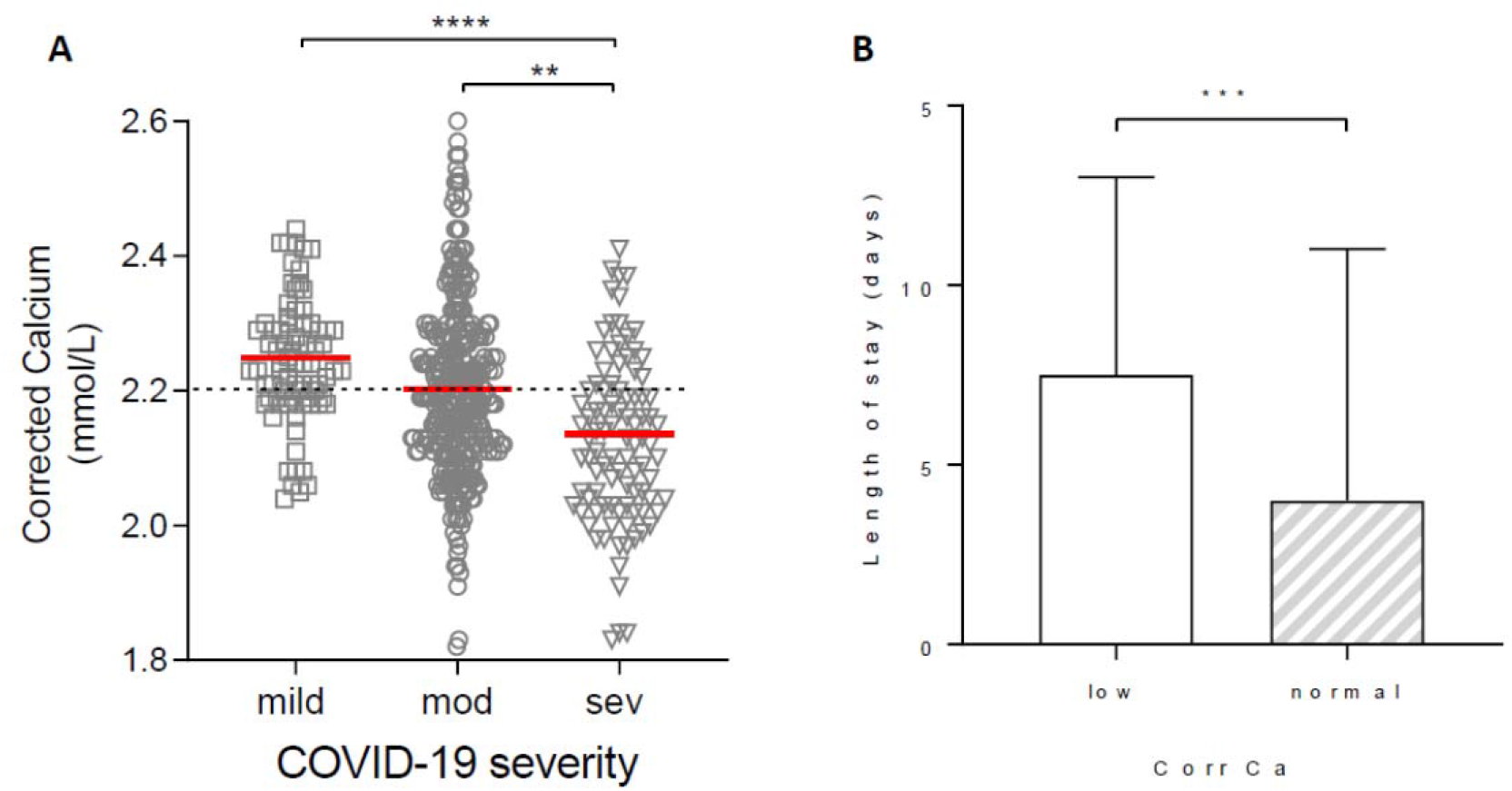
Association of calcium levels in patients with COVID-19 and disease severity or length of stay (LOS) (A) Calcium levels in patients with mild disease (no supplemental oxygen requirement managed in a virtual hospital setting), moderate disease (patients requiring admission to hospital), severe disease (patients requiring additional respiratory support including invasive and non-invasive ventilation) (B) LOS in hypocalcaemic (corrected calcium < 2.2mmol/L) and normocalcaemic (≥ 2.2mmol/L) COVID-19 patients. Plots show median ± IQR ** p<0.01 *** p< 0.001 **** p<0.0001

Hypocalcaemia was more common in patients with COVID-19 segregated by increasing level of hospital care. Whereas 27.8% (22/79) of patients with mild ambulatory COVID-19 were hypocalcaemic, the rate doubled to 53.3% (175/328) among those with moderate severity COVID-19. The proportion was higher still (72.1%; 70/97) in individuals with severe COVID-19 (**Table 2**). Hypocalcaemia at presentation was thus associated with a higher unadjusted odds ratio for severe COVID-19 (2.74; 95% CI 1.69-4.43) As expected, hypocalcaemia was also associated with an increased length of hospital stay compared to patients who had normal corrected calcium (median 7.5 vs 4.0 days; p<0.0001) (**Figure 3B**). In contrast, hypocalcaemia in patients with CAP and VP was not associated with either disease severity or length of stay **(Supplementary Figure 1 and Supplementary Table 1)**.

**Table 2.**
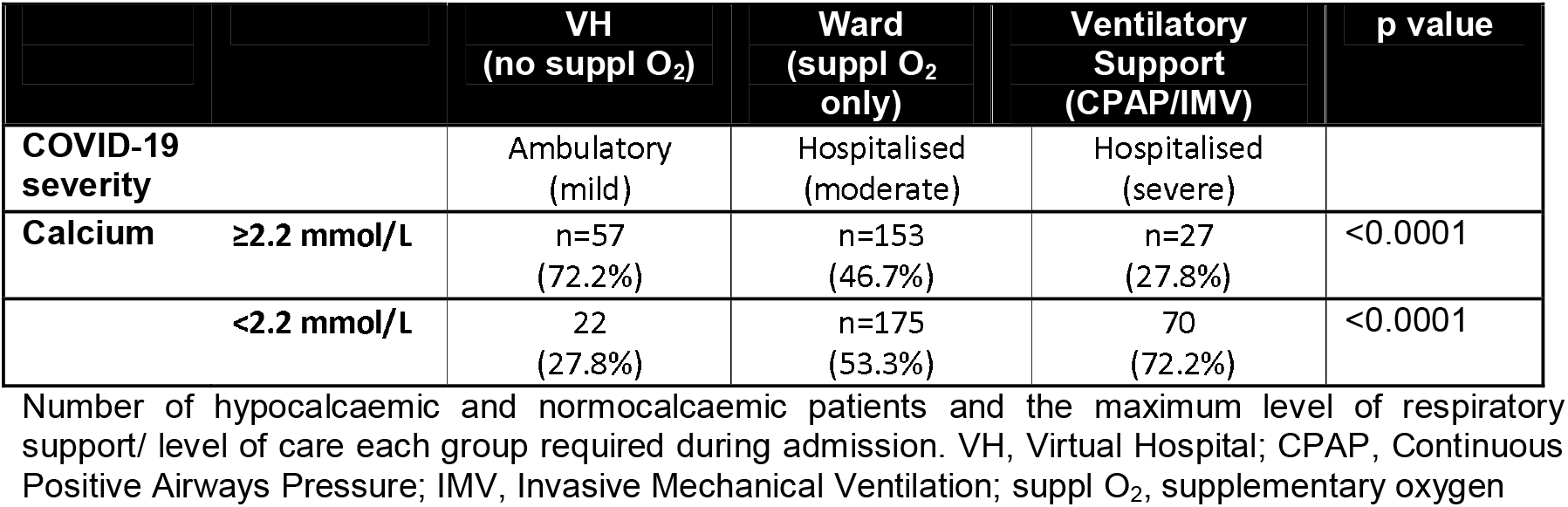
Hypocalcaemia in COVID-19 patients at different levels of care.

Evaluation of hypocalcaemia as an independent predictor of a higher level of clinical care, a practical surrogate for requirement for higher respiratory support, was undertaken using ordinal logistic regression. In recognising that COVID-19 pneumonia is associated with a wide range of severity and that patients may progress from initially mild to more consequential disease, our analyses made two assumptions: first, that the outcome categories (different care levels) followed a natural order within the studied range of disease severity, and second, proportional odds assumption of the effect of hypocalcaemia on the same outcome categories was obeyed. A goodness-of-fit test, in this case the Brant test, was used to determine proportional odds assumption (**Supplementary table 2**).

Based on initial analyses, a partial proportional odds model was developed that excluded increasing age as an exploratory variable as it violated the proportional (or parallel) odds assumption rule for the chosen outcome (Peterson and Harrell, 1990). Older age has been shown in numerous studies to be the most dominant independent predictor of death in COVID-19, suggesting that its prognostic impact on any categorised COVID-19 outcome was likely to be disproportionate. An analysis of ordinal logistic regression was thus formulated based on maximum likelihood fit of the predictive model to the data (**Table 3**). In the final model, variables with the highest odds ratios (all ≥2.00) for predicting a higher level of care were chest radiographic zones, hypocalcaemia, S:F ratio and CRP.

**Table 3.** Ordinal Regression Analysis.

| Variable |  | OR | 95% CI | p value |
| --- | --- | --- | --- | --- |
| CXR zones | >4 abnormal | 2.48 | 1.53-4.01 | <0.001 |
| Calcium | <2.2mmol/L | 2.33 | 1.50-3.61 | <0.001 |
| S:F ratio | <4.38 | 2.16 | 1.33-3.52 | 0.002 |
| CRP | >50mg/L | 2.00 | 1.23-3.25 | 0.005 |
| Respiratory Rate | >24/min | 1.67 | 1.06-2.65 | 0.028 |
| Leucocytosis | >11 x 10<sup>9</sup>/L | 1.37 | 0.80-2.32 | 0.252 |
| Lymphopenia | <0.7 x 10<sup>9</sup>/L | 0.96 | 0.62-1.50 | 0.872 |

Partial Proportional Odds Model that shows the odds ratio (OR) of each variable in predicting the need for higher level of care in COVID-19 pneumonia. S:F ratio= saturations: fraction of inspired oxygen ratio.

### Normalisation of serum calcium level in survivors of severe COVID-19

To assess trends in calcium level over the course of COVID-19, we identified two groups for analysis: survivors (n=74; 56% male; median age 76, IQR 60 - 80) and non-survivors (n=60; 61% male; median age 79, IQR 62-83). Both groups shared a similar level of baseline hypocalcaemia (2.12 mmol/L vs 2.13 mmol/L respectively), median length of hospital stay (11 vs 10 days) and avoidance of ICU. Survivors and non-survivors remained hypocalcaemic until day 4-6, though there was a significant difference between the two groups (2.18 mmol/L versus 2.13 mmol/L respectively, p=0.02). By day 7-9, serum calcium had normalised to 2.25 mmol/L amongst survivors whereas it remained below the reference range 2.13 mmol/L in non-survivors, similar to admission (p< 0.0001). The difference in corrected calcium concentration was even greater 9 days or more after admission. At discharge from hospital, the median calcium concentration was 2.3 mmol/L in survivors versus 2.15 mmol/L in non-survivors (**Figure 4**). None of these patients received calcium supplementation during their hospital admission.

**Figure 4.**
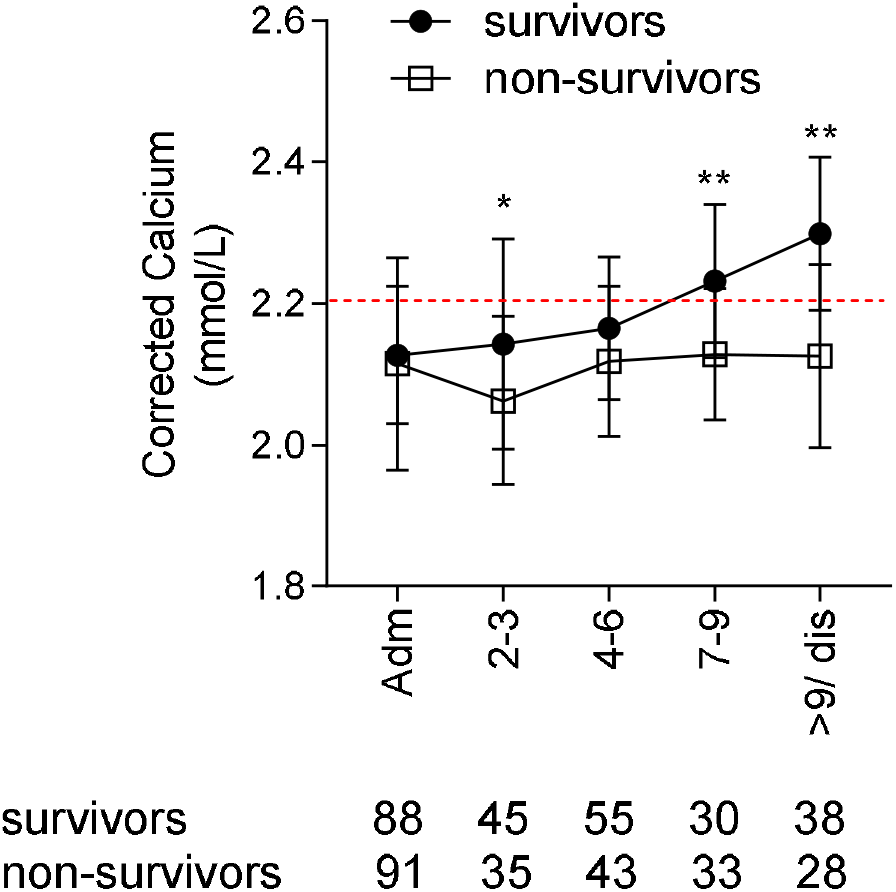
Corrected calcium levels during admission in survivors and non-survivors of COVID-19, hypocalcaemic at presentation. Calcium levels compared at each time point. Plots show median and IQR, * p value <0.05 ** p value < 0.01

## Discussion

In this retrospective analysis, we found hypocalcaemia to be a common feature of COVID-19, with a frequency and degree of derangement that was greater than in non-COVID-19 community-acquired pneumonia and viral pneumonia. We also observed that low calcium in COVID-19 was associated with measures of abnormal physiology including oxygenation and respiratory rate, radiographic extent of pneumonic involvement and elevated CRP.

Low calcium was specifically linked to severe disease and different levels of hospital care aligned with increasing levels respiratory support (a surrogate index of severity of COVID-19 pneumonia). Ordinal regression of risk estimates for categorised care levels showed that baseline hypocalcaemia was incrementally associated with an over two-fold risk for a higher level of care. Its adjusted odds ratio (2.33) was higher than that for variables that have previously been shown to predict worse COVID-19 outcome, such as hypoxia and tachypnoea. It was also greater than the odds ratio attributed to CRP and nearly as high as that for the extent of radiographic disease.

Unlike some of these parameters, hypocalcaemia has not been included in COVID-19 severity prediction scores (2,3,23). This could be because advancing age and CKD are predictors of poor outcome in COVID-19, both of which impact calcium homeostasis. Older adults are also more likely to be on calcium supplementation. However, the distinct association between baseline hypocalcaemia and COVID-19 severity suggests that it could potentially contribute to multivariable stratification of COVID-19 outcome, and importantly, predict the need for additional respiratory support in a select subgroup of young patients without CKD.

Hypocalcaemia has previously been associated with a higher risk of hospitalization (5), ICU admission (10,12), ventilation and, in some cases, mortality in patients with COVID-19 (8). Our data is consistent with these findings. However, the rate of hypocalcaemia in our study of 53% was lower than 62 to 74.4% reported in other studies (4,5,7–10)). This difference may be due to case-mix or cohort heterogeneity, specifically the inclusion of patients with mild disease who were managed in a virtual hospital setting in our study.

Low serum calcium is most commonly caused by Vitamin D deficiency, insufficient PTH production and renal dysfunction. In order to reduce the confounding effect of renal failure on hypocalcaemia, these patients were excluded from analysis. There is conflicting evidence on the link between Vitamin D deficiency and COVID-19 severity; one retrospective study from Israel reported independent associations between low pre-pandemic Vitamin D levels and subsequent incidence and severity of COVID-19 (24), but an analogous study in the UK failed to show such associations (25). In a preliminary analysis of 30 patients with mild versus severe COVID-19, we found that whereas calcium levels were significantly lower in severe disease, Vitamin D levels were not (Supplementary Figure 2). A larger sample size, matched for age and ethnicity (which both impact Vitamin D levels) is necessary to further corroborate these preliminary findings.

The role of PTH in COVID-19 is unclear but normal PTH levels have been reported in a limited study of hospitalized patients with this disease (10). Whilst it is unlikely that PTH insufficiency is the sole or even primary driver of hypocalcaemia in COVID-19, this area requires further investigation.

Two clinically important coronaviruses, SARS-CoV-1 and Middle East respiratory syndrome (MERS)-CoV, can use calcium ions to enter host cells, mediated by a fusion peptide derived from the viral spike protein (26,27). It could be postulated that similar calcium-dependent mechanisms might operate during the pathogenic process involving calcium handling during SARS-CoV-2-mediated infection. In support of this hypothesis, calcium channel blockers including amlodipine have been shown to inhibit post-entry replication events of SARS-CoV-2 in vitro (28). In one observational study, amlodipine therapy was also associated with a decreased case fatality rate in hospitalized COVID-19 patients (28). Other mechanisms of calcium-mediated inflammation and tissue injury may involve the activation of the viral NLRP3 inflammasome leading to IL-1beta over-production and downstream activation and propagation of tissue inflammatory cascades (Nieto-Torres 2015; Cron, 2021; Huet et al., 2020; Webb et al., 2020).

Hypocalcaemia is common in critically ill patients, with a reported prevalence of up to 88% of all patients admitted to intensive care (29,30). In our study, baseline hypocalcaemia was frequently evident in COVID-19 patients even amongst those who were not ultimately admitted to the ICU. Patients with COVID-19 also had comparable baseline illness characteristics as CAP, such as respiratory rate, S:F ratio, prevalence of elevated CRP, frequency of lymphopenia. However, the frequency of hypocalcaemia was significantly higher and the median concentration of serum calcium lower in those with COVID-19 patients compared to those with CAP. This finding suggests the possibility of hypocalcaemia being pathophysiologically more pertinent in COVID-19. In line with this, a lower serum calcium level within the COVID-19 subgroup was clearly associated with worse respiratory illness and a need for greater respiratory support.

Our study was limited by its retrospective design and resultant occurrence of missing data (<15% for COVID-19 and CAP subgroups), with the fullest data available for COVID-19 and the least for patients with VP. The modest sample size of our COVID-

19 cohort was dependent on the incident caseload during the first wave of the pandemic, and a small sample size of CAP and VP patients. The data on vitamin D and PTH measurements was insufficient as these parameters were not routinely measured in the management of COVID-19 or the other pneumonias. In addition the measurement of ionized calcium (the unbound calcium available to cells) would be more accurate and physiologically relevant than measuring the total body calcium by corrected for albumin (31). However, in the absence of ionized calcium measurements, corrected calcium was felt to be a reasonable alternative that more accurately reflected calcium stores compared to calcium that was not adjusted for protein binding.

Our data show that hypocalcaemia is clinically important during both the acute presentation of COVID-19 as well as its subsequent evolution to more severe respiratory failure. Further studies are required to help bridge the biological understanding of hypocalcaemia and its pathophysiological significance to this devastating disease. If calcium is ultimately shown to be critical to the entry and replication of SARS-CoV-2 in host cells, unravelling how this mechanism could be therapeutically targeted deserves more intensive examination.

## Data Availability

Data are available upon reasonable request. Deidentified participant data may be requested from the corresponding author following publication of the study.

## Declarations

### Ethics approval and consent to participate

Ethical approval was granted for the prospective recruitment of human data on adult patients with positive SARS-CoV-2 nucleic acid testing by real-time polymerase chain reaction (rRT-PCR) at Watford General Hospital, West Hertfordshire NHS Trust (NHS HRA: 20/HRA/2344).

### Consent for publication

Not applicable

### Availability of data and materials

The datasets generated and/or analysed during the current study are not publicly available as individual privacy would be compromised, but are available from the corresponding author in anonymized format, on reasonable request.

### Author’s contributions

Study Concept and Design: MM, AB and RV. Acquisition of data MM, HG, SC, MP, RS, TV. Analysis and Interpretation of Data: MM, FC, AD, RV. Drafting of the manuscript: MM, FC, AD, RV. Critical revision of manuscript: All authors

## Acknowledgements

Not applicable

## Supplemental Figure Legends

Supplemental Figure 1 **Calcium levels in patients with COVID-19, CAP or VP according to disease severity**. Calcium levels at presentation in patients with mild, moderate and severe disease as defined for each subgroup (see methods). Red line represents median, ns= not significant, ** p<0.01, **** p<0.0001

Supplemental Figure 2 **Paired calcium and Vitamin D levels in patients with mild versus severe COVID-19 disease**. Calcium levels at presentation, and paired Vitamin D levels measured within three days of a positive SARS-CoV-2 PCR positive result. Red line represents median, ns= not significant, ** p<0.01,

## Supplemental Tables

**Supplementary Table 1.** LOS in hypocalcaemic patients with non-COVID pneumonia

|  | Community Acquired Pneumonia |  |  | Viral Pneumonia |  |  |
| --- | --- | --- | --- | --- | --- | --- |
| Calcium | Low | Normal | P value | Low | Normal | P value |
| Median LOS (days) | 6 | 5 | 0.74 | 4 | 4 | 0.47 |

LOS in hypocalcaemic (corrected calcium $< 2.2\text{mmol/L}$ ) and normocalcaemic ( $\geq 2.2\text{mmol/L}$ ) patients with Community Acquired Pneumonia (CAP) and Viral Pneumonia (VP). LOS= length of stay

**Supplementary Table 2.** Brant test for goodness-of-fit prior to ordinal logistic regression

|  | Chi<sup>2</sup> | P value | Df |
| --- | --- | --- | --- |
| Age | 36.3 | $< 0.001$ | 2 |
| CXR zone ($\geq 4$ abnormal) | 3.33 | 0.190 | 2 |
| Lymphopenia ($< 0.7 \times 10^9/\text{L}$) | 0.84 | 0.657 | 2 |
| CRP ($> 50 \text{ mg/L}$) | 0.44 | 0.805 | 2 |
| Leucocytosis ($> 11 \times 10^9/\text{L}$) | 0.49 | 0.782 | 2 |
| Calcium (<2.2 mmol/L) | 3.50 | 0.174 | 2 |
| Respiratory rate (>24/min) | 1.34 | 0.511 | 2 |
| S:F ratio (define) | 2.96 | 0.227 | 2 |

A significant test statistic ( $P < 0.05$ ) is evident that the proportional odds assumption has been violated. Df, degrees of freedom.

