## Supplementary figures and images for "Increased prevalence and clinical impact of hypocalcaemia in severe COVID-19 distinguishes it from other forms of infective pneumonia"

### Supplemental Figure 1

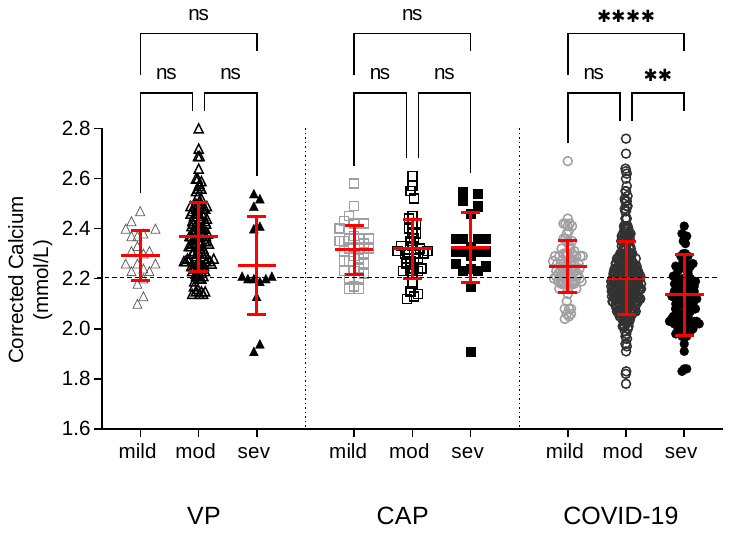

### Supplemental Figure 2

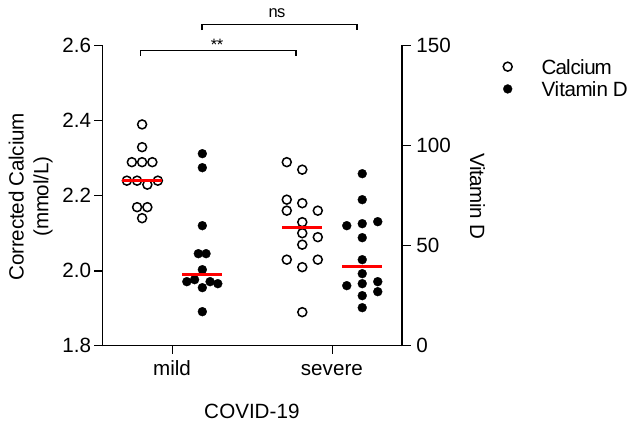
